# Diagnostic agreement and accuracy of dermatopathology versus molecular PCR test in distinguishing eczema from psoriasis

**DOI:** 10.1101/2025.10.29.25339084

**Authors:** A Andrea Schmitt, S Proksch, Ludwig Gutzweiler, Sandra Roth, Marcella Engler, Cornelia S. L. Müller, Andreas Volz, Andreas W. Arnold, Monika Šedivcová, Adriana Bernklauova, Miroslav Dura, Denisa Kacerovska, Katja Technau-Ihling, Christian Ihling, Christiane Rakozy, Wiebke Pruessmann, Thomas Leibing, Maria Isabel von Eichborn, Johannes Kern, Elisabeth Oms, Stefanie Eyerich, Kilian Eyerich, Helmut Laaff, Natalie Garzorz-Stark, Kristin Technau-Hafsi

## Abstract

**Background:** Targeted treatments for non-communicable chronic inflammatory skin diseases like eczema and psoriasis offer significant potential for effective therapy. However, therapeutic success requires an accurate diagnosis, which is challenging due to their overlapping clinical and histological features.

**Objective:** We aimed at assessing the diagnostic performance of both a manual (MC) and fully automated (PsorX-LabDisk) RT-qPCR test based on the expression of *NOS2* and *CCL27* compared with conventional dermatopathological evaluation in differentiating psoriasis from eczema.

**Methods:** Seventy-three FFPE skin samples of psoriasis and eczema were randomly selected and evaluated histopathologically (H&E-stained sections) by 14 dermatopathologists to assess interobserver variability, quantified using Cohen’s and Fleiss’ κ. To confirm that the observed variability was not cohort- or rater-specific, a validation cohort (n=72) from an independent institution was assessed by three dermatopathologists under identical conditions. For molecular analysis, both manual (MC) and automated *NOS2/CCL27*-based RT-qPCR (PsorX-LabDisk) workflows were applied. Diagnostic performance (sensitivity, specificity, accuracy) of histopathological and molecular analyses were determined against reference diagnoses.

**Results:** Dermatopathological evaluation demonstrated only fair agreement (Fleiss’ κ = 0.31) in both study and validation cohort. The mean diagnostic accuracy of dermatopathology was 76.9%, with a sensitivity of 70% and specificity of 81.6%. In comparison, MC and the PsorX-LabDisk achieved sensitivities of both 92.9%, specificities of 82.2% and 84.4%, and accuracies of 87.7% and 86.3%, respectively. In diagnostically ambiguous cases, molecular testing maintained high accuracy (>86%), clearly outperforming dermatopathology, which showed near-random agreement and low accuracy (61.7%).

**Conclusions:** Both MC and PsorX-LabDisk provide a reliable, examiner-independent complement to dermatopathology for differentiating psoriasis and eczema. By reducing diagnostic ambiguity, it enhances clinical confidence and supports more precise and timely therapeutic decisions in inflammatory skin disease management.

**Key points:** *High interobserver variability in dermatopathology:* Across two independent cohorts, dermatopathological evaluation by multiple dermatopathologists showed only fair to no agreement, highlighting substantial subjectivity and diagnostic uncertainty in distinguishing psoriasis from eczema based solely on morphology.

*Superior accuracy of molecular diagnostics:* Both the manual (MC) and fully automated *NOS2/CCL27*-based RT-qPCR (PsorX-LabDisk) assays outperformed dermatopathology, achieving sensitivities around 93 % and overall accuracies around 88 %, demonstrating that molecular testing provides a more consistent and objective diagnostic approach.

*Robust performance in ambiguous cases:* In diagnostically challenging samples with low dermatopathological consensus, the PsorX-LabDisk maintained high diagnostic accuracy (>86 %), outperforming expert evaluation. These results underscore its potential as a reliable, examiner-independent tool supporting precise diagnosis and optimized treatment selection in clinical practice.

**Capsule Summary:** Both MC and PsorX-LabDisk molecular assay outperformed dermatopathology in differentiating psoriasis from eczema, offering an objective, reproducible, and clinically practical tool that enhances diagnostic confidence and guides targeted treatment in inflammatory skin diseases.

## Introduction

Inflammatory skin diseases such as psoriasis and eczema are among the most prevalent chronic dermatologic conditions worldwide, with considerable implications for patient quality of life and healthcare resource utilization. Psoriasis affects 2–3% of the global population, whereas atopic dermatitis (AD) is reported in up to 20% of children and 10% of adults, with rising prevalence in industrialized countries (*1–3*). While clinical presentation remains central to diagnosis, dermatopathological evaluation is frequently employed to confirm or differentiate these entities, particularly in atypical or treatment-refractory cases (*4, 5*).

Yet, even at the histomorphological level, psoriasis and eczema can share features such as epidermal hyperplasia, parakeratosis, and lymphocytic infiltrates, complicating reliable distinction (*6*). As dermatopathology is qualitative and inherently subjective, interobserver variance is to be expected. Studies on inter-observer agreement in chronic inflammatory skin diseases have largely addressed clinical severity scoring - such as PASI for psoriasis and EASI or SCORAD for AD (*7–9*) - with intra- and interrater reliability ranging from fair to good, whereas comparable data for pathologist concordance in biopsy interpretation for these diseases are lacking. The absence of such data is notable given the central role of dermatopathology in the diagnostic process and thus its impact on therapeutic decision-making. The growing repertoire of biologics targeting cytokines such as TNF, IL-17, IL-23, and IL-4/13 has significantly improved outcomes in moderate-to-severe psoriasis and AD, but also raised the stakes for accurate diagnostic classification (*6, 10–12*). Misclassification of a lesion - for instance, interpreting dermatitis as psoriasis or vice versa - may lead to inappropriate therapy, delayed access to effective biologics, or exposure to unnecessary systemic immunosuppressants, all of which have implications for patient safety and public health costs (*13, 14*). Several strategies have been proposed to improve diagnostic consistency in inflammatory dermatoses including digital pathology platforms with AI-assisted pattern recognition and the use of immunohistochemical adjuncts such as IL-36γ to support or refine morphologic diagnoses (*15, 16*). Importantly, molecular diagnostics are increasingly recognized as a promising advancement in dermatopathology, particularly for differentiating between psoriasis and eczema in histologically ambiguous cases. Quaranta et al. showed that psoriasis and eczema exhibit distinct immune signatures, with psoriasis characterized by a Th17/IL-17/IL-36 profile and eczema by a Th2/IL-4/IL-13 signature with barrier dysfunction. On this basis, a molecular classifier using *NOS2* and *CCL27* gene expression was developed that accurately distinguished between the two diseases, including clinically ambiguous or initially misclassified cases (*17, 18*). Moreover, the assay proved effective in FFPE tissue, underscoring its suitability as molecular tool for routine diagnostics (*14*). The present study addresses two key questions: How reproducible is dermatopathological differentiation of psoriasis and eczema among dermatopathologists? And can both a manual and a fully automated qPCR assay - available as the PsorX -LabDisk (Dermagnostix, Germany) enabling rapid, standardized and fully automated analysis of FFPE biopsies - enhance diagnostic accuracy compared with conventional pathology? By evaluating inter-observer variability and benchmarking molecular against dermatopathological performance, we aimed to define the limitations of morphology and examine the added value of molecular diagnostics in routine dermatopathology.

## Material and Methods

### Patient cohorts

This retrospective multi-center study included two cohorts from Germany (Study cohort) and the Czech Republic (Validation cohort). The study cohort comprised 73 FFPE archived skin biopsy samples randomly selected at the Department of Dermatology, Medical Center Freiburg, from patients with diagnosed psoriasis or eczema (n= 28 psoriasis, n= 45 eczema). The validation cohort was used to verify that interrater variability patterns were reproducible across raters and samples and comprised 72 archived FFPE skin samples collected at Biopticka Laborator (Pilsen, Czech Republic) from patients with clinically suspected psoriasis or eczema. The reference diagnosis of all patients in the study cohort was determined by three dermatologists based on all available metadata in a tertiary care university hospital including initial clinical suspicion, clinical picture, family history, laboratory parameters, course of disease including therapeutic outcome, and dermatopathology report. In the validation cohort, a final reference diagnosis could not be established due to limited clinical information as expected for a referral laboratory. All patients gave written informed consent, and the study was approved by the local ethical committee of the University of Freiburg (project number 25-1009-S1) and the Faculty of Medicine in Pilsen (project number 122/25). Further patient characteristics are listed in Table 1 and 2.

**Table 1:**
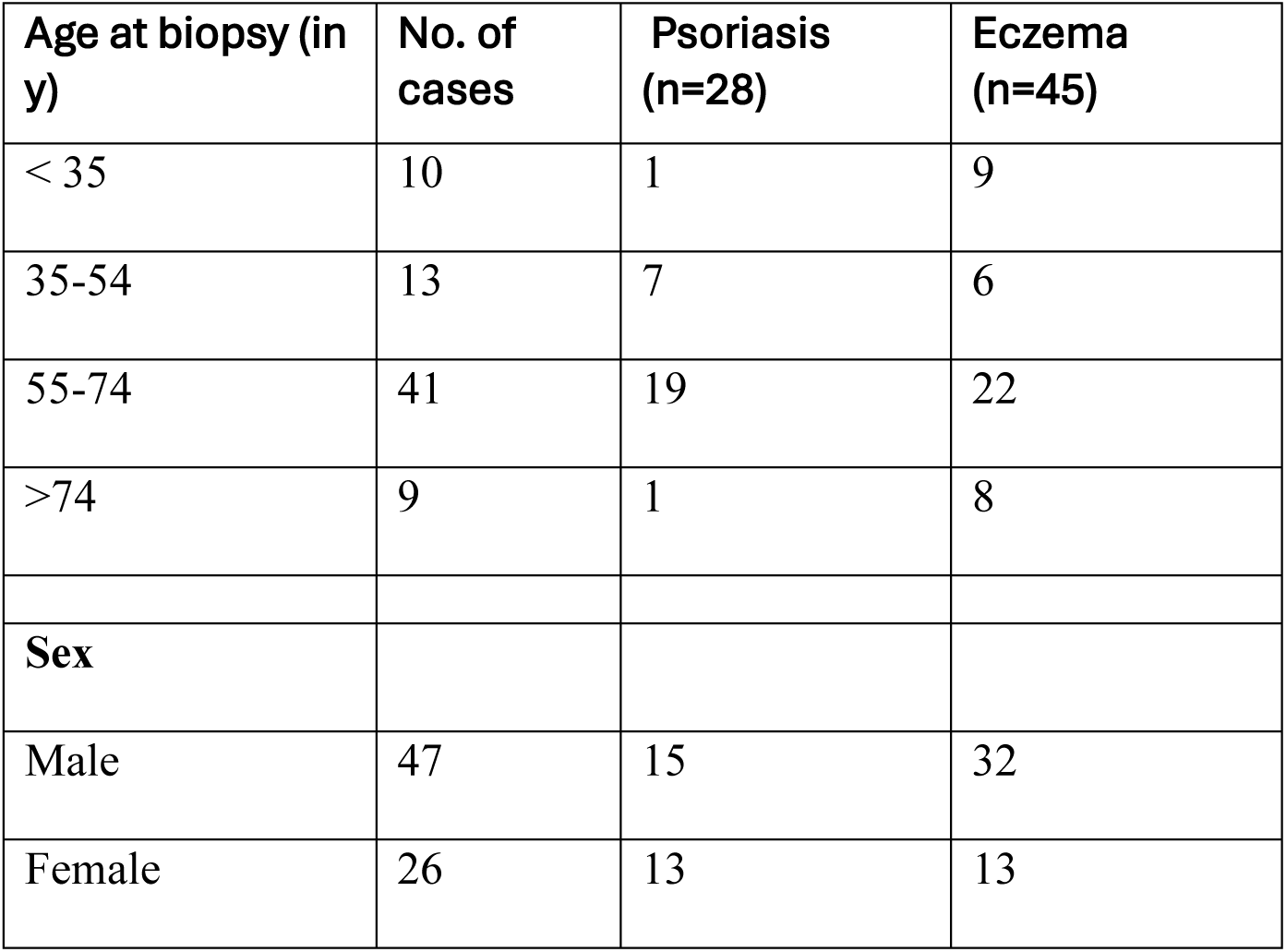
Patient Characteristics of the study cohort.

**Table 2:** Patient Characteristics of the validation cohort.

| Age at biopsy (in y) | No. of cases with psoriasis/eczema |
| --- | --- |
| < 35 | 17 |
| 35-54 | 30 |
| 55-74 | 19 |
| >74 | 6 |
| **Sex** |  |
| Male | 39 |
| Female | 33 |

### Dermatopathological assessments for analysis of interobserver reliability

In the study cohort, 73 samples were randomly selected, and their corresponding H&E-stained slides were evaluated by 14 independent board-certified dermatopathologists. The validation cohort included H&E-stained slides of 72 skin samples, which were assessed by three independent board-certified dermatopathologists. All evaluations were performed in a blinded manner, and the observers were unaware of each other’s assessments to minimize bias. The original clinical suspicion was provided to all evaluators. Dermatopathological classification was performed using six categories: eczema, psoriasiform eczema, undetermined, eczematized psoriasis, psoriasis, and diagnosis other than psoriasis or eczema. For subsequent analyses, these categories were consolidated into three groups: psoriasis, eczema, and undetermined.

### Molecular assessment and comparison of molecular classifier result to reference diagnosis

Molecular testing was performed on formalin-fixed, paraffin-embedded (FFPE) skin biopsy specimens using two parallel workflows in the study cohort, namely manual quantitative real-time PCR (qPCR) and a fully automated PCR-based diagnostic platform (Dermagnostix LabDisk-Analyer and PsorX-LabDisk). Manual PCR was performed following the workflow, primers, and probes previously described by Fischer et al. (*19*) to ensure comparability with published protocols. Briefly, real-time PCR assays for FFPE samples were carried out in 96-well plates using a one-step RT-PCR master mix (Thermo Fisher Scientific, Waltham, MA, USA). Thermocycling and fluorescence detection were conducted on a qTower³ system (Analytik Jena, Germany), and amplification data were processed using qPCRsoft 4.1 (Analytik Jena). Expression levels of the target genes *NOS2* and *CCL27* were normalized to reference genes *TBP* and *SDHAF2* and analyzed using the previously described algorithm (*19*). In parallel, the same FFPE samples were analyzed using a fully automated, cartridge-based PCR system (PsorX-LabDisk, Dermagnostix, Germany), which integrates RNA extraction, purification, reverse transcription, amplification, and analysis into a single workflow. Results of PsorX-LabDisk were available within approximately two hours. Both the manual and automated PCR workflows generated an algorithm-derived probability for each sample, classifying it as “psoriasis probable” (probability > 55%), “undetermined” (≥ 45% and ≤ 55%), or “psoriasis improbable” (probability < 45%).

### Statistical Analysis

All statistical analyses were performed using Microsoft Excel and Python 3 (Spyder 6 IDE), employing the following libraries: pandas, numpy, and scikit-learn. Inter-observer agreement and diagnostic performance were assessed using established statistical methods. Cohen’s kappa (κ) was applied to quantify pairwise agreement between two raters beyond chance. Fleiss’ kappa (κ) was used to measure multi-rater agreement among three or more observers, providing a generalized measure of concordance across multiple evaluators. Both statistics account for agreement expected by chance and are interpreted according to Landis and Koch: κ < 0 indicates poor, 0-0.20 indicates slight, 0.21–0.40 fair, 0.41–0.60 moderate, 0.61–0.80 substantial, and 0.81–1.00 almost perfect agreement. Samples were categorized as “ambiguous” when diagnostic consensus of raters was low, defined as concordance among fewer than 10 of 14 dermatopathologists in the study cohort or disagreement by at least one of three raters in the validation cohort. Diagnostic performance of the molecular classifier relative to the reference diagnosis was assessed by calculating sensitivity and specificity using a fourfold contingency table.

## Results

### Dermatopathological assessment of psoriasis and eczema cases revealed substantial inter-observer variability

Seventy-three FFPE samples from psoriasis and eczema patients were evaluated by 14 independent dermatopathologists and assigned to psoriasis, eczema, undetermined or other diagnosis (Figure 1A). 38% of samples corresponded to the reference diagnosis of psoriasis (n = 28) and 62% to the reference diagnosis of eczema (n = 45). The overall distribution of diagnostic categories assigned by the individual raters roughly reflected the proportions of the reference diagnoses, indicating no systematic bias toward either psoriasis or eczema in the study cohort (Fig. 1B). However, when assessed at the level of individual samples, a considerable number of false-positive and false-negative classifications became evident (Fig. 1C). Eczema tended to be diagnosed correctly more frequently than psoriasis (82% ± 6.7% for eczema, and 70% ± 13.7% for psoriasis), psoriasis cases were more often underdiagnosed (mean false negative rate 30% ± 13.7%) than eczema (mean false positive rate 18% ± 6.7%), with variable true-positive and false-positive rates across raters. Pairwise comparison of diagnostic performance independent of the reference diagnosis revealed Cohen’s κ values ranging from only 0.12 to 0.62 (Fig. 1D). To assess if low interrater consistency is a general phenomenon, we had three different pathologists evaluate a different cohort of n=72 psoriasis and eczema samples in an identical experimental setting. Also in this cohort, Cohen’s κ values ranged from 0.32 to 0.67 (Fig. 1E). Overall inter-observer agreement, as determined by Fleiss’ κ, was fair to moderate (κ = 0.31 in both cohort 1 and 2), reflecting the intrinsic heterogeneity of dermatopathological interpretation in inflammatory skin diseases (Fig. 1F).

**Figure 1:**
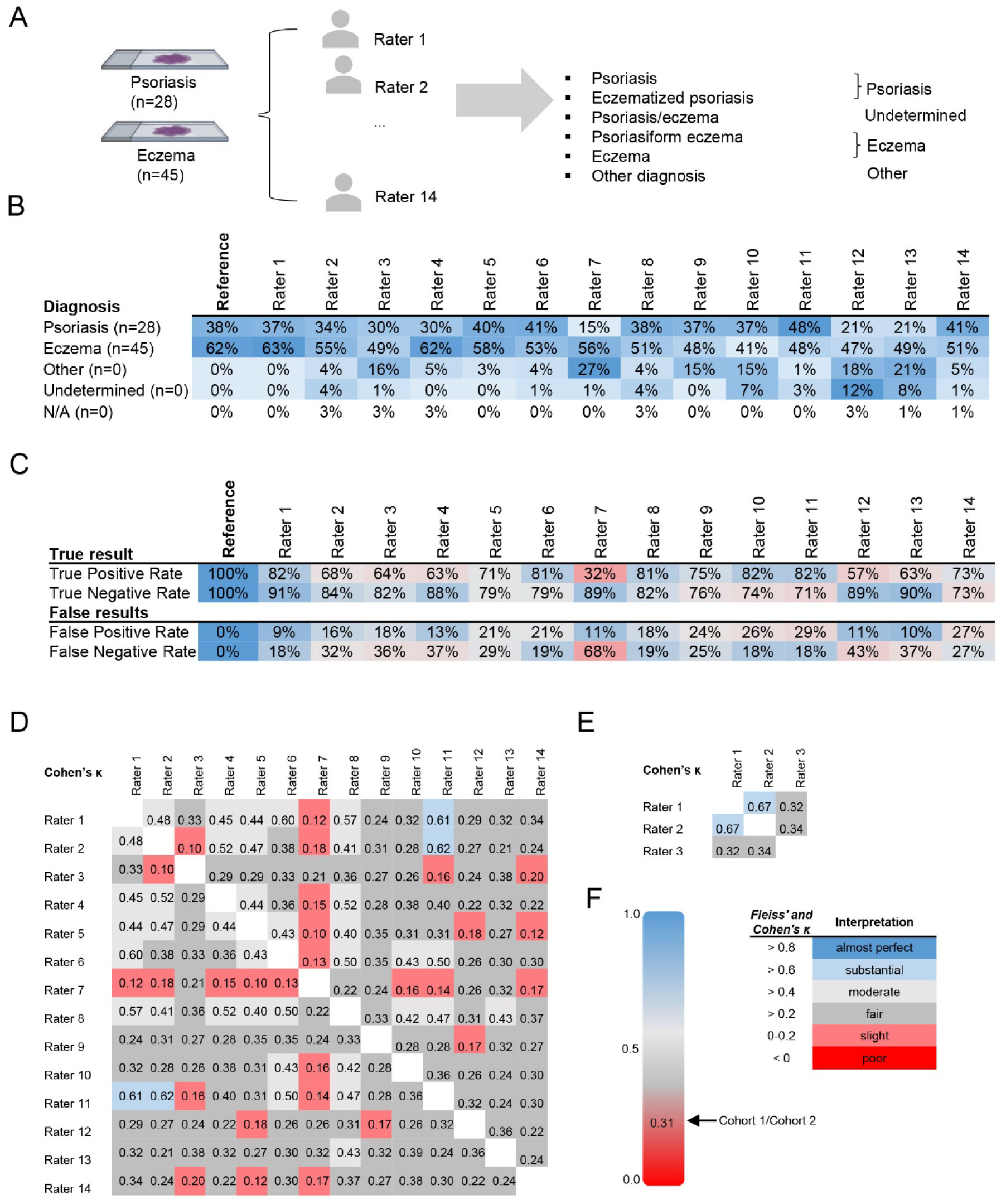
Interrater variability and diagnostic performance among dermatopathologists. **(A)** Study design: skin biopsies from patients with psoriasis (n = 28) and eczema (n = 45) were independently evaluated by 14 dermatopathologists. Each rater assigned one of the following diagnoses: psoriasis, eczematized psoriasis, psoriasis/eczema, psoriasiform eczema, eczema, or other diagnosis. For analysis, results were grouped into *psoriasis*, *eczema*, *undetermined*, or *other*. **(B)** Distribution of diagnoses assigned by 14 dermatopathologists (Rater 1-14) compared to the reference diagnosis in the study cohort. **(C)** Diagnostic performance metrics for each rater. True positive rate (TPR) = proportion of psoriasis cases correctly identified as psoriasis; true negative rate (TNR) = proportion of eczema cases correctly identified as eczema; false positive rate (FPR) = proportion of eczema cases incorrectly diagnosed as psoriasis; false negative rate (FNR) = proportion of psoriasis cases incorrectly diagnosed as eczema. **(D-E)** Pairwise interrater agreement assessed using Cohen’s κ for the study cohort (D) and the validation cohort (E). **(F)** Agreement as measured by Fleiss’ κ and Cohen’s κ with interpretation according to Landis & Koch.

### Both manual molecular classifier and automated PsorX-LabDisk shows robust diagnostic performance

We next aimed to evaluate the diagnostic accuracy of each dermatopathologist by calculating sensitivity for the correct identification of psoriasis and specificity for the correct exclusion of psoriasis (i.e. correct diagnosis of eczema) in comparison with the reference diagnosis in the study cohort (Fig. 2A). Sensitivity values varied substantially among dermatopathologists, ranging from 32% to 82.1%, with an overall sensitivity of 70%. Specificity values were more consistent, ranging from 71.4% to 91.1% (mean specificity = 81.6%), corresponding to an overall diagnostic accuracy of 76.9% (Fig. 2B and C). Given the high inter-observer variability, we evaluated the performance of the molecular assay designed to distinguish psoriasis from eczema using both a commercially available fully automated system (PsorX-LabDisk) and a manual qPCR workflow as reference method in the study cohort. PsorX-LabDisk is a cartridge-based molecular diagnostic assay built on a centrifugal microfluidics (lab-on-a-disk) platform, enabling fully automated sample processing from input to readout. The system integrates modules for sample preparation, nucleic acid purification, amplification (RT-PCR), and detection, using a mix of lyophilized reagents and liquid reagents housed in stick-packs allowing hands-off operation (Fig. 2A). Sensitivity of the conventional manual PCR workflow was 92.9%, specificity was 82.2% and accuracy was 86.3%. Using the conventional manual PCR workflow as benchmark, the positive percent agreement sensitivity of PsorX-LabDisk was 94.1%, negative percent agreement was 97.4% and overall percent agreement was 95.9% compared to the manual PCR, showing that the fully automated mode of operation sustains the high precision of molecular testing. More importantly, PosrX-LabDisk demonstrated high and consistent diagnostic performance when compared to the reference diagnoses, with a sensitivity of 92.9%, specificity of 84.4%, and overall accuracy of 87.7% (Fig 2B and C).

**Figure 2:**
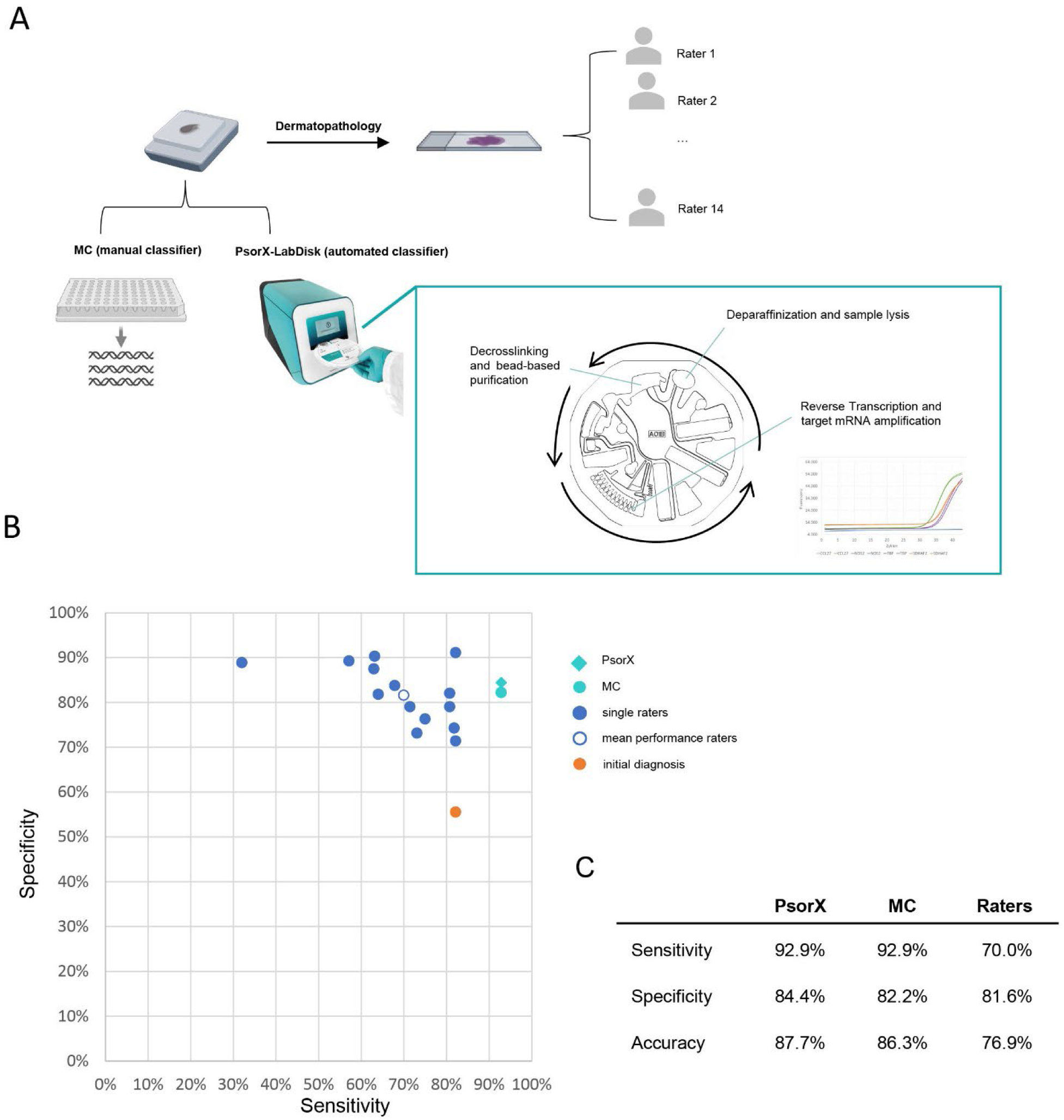
Diagnostic performance of the molecular classifier (MC), automated PsorX-LabDisk system, and dermatopathologists. **(A)** Performance Evaluation of Raters, Manual Classifier, and PsorX-LabDisk. PsorX-LabDisk (in turquoise box) is a single-use disposable microfluidic cartridge enabling fully automated processing of formalin-fixed, paraffin-embedded (FFPE) tissue samples. The integrated workflow, including automated deparaffinization and sample lysis, decrosslinking and bead-based nucleic acid purification, reverse transcription, and target mRNA amplification are schematically shown. The system performs all steps from sample input to qPCR readout without manual intervention, enabling standardized, hands-free molecular diagnostics. **(B)** Sensitivity and specificity of individual dermatopathologists (blue dots), mean performance of all dermatopathologists (white dot outlined in blue), initial clinical diagnosis (orange), and molecular assays (MC, turquoise; PsorX, turquoise square) in the study cohort. **(C)** Summary of diagnostic performance metrics showing sensitivity, specificity, and overall accuracy for the manual molecular classifier (MC), fully automated PsorX-LabDisk system, and dermatopathological raters in the study cohort.

### Molecular classification clearly outperforms dermatopathologists in ambiguous cases

The reference diagnosis matched the initial clinical suspicion in only 48 of 73 cases (65.7%) suggesting inclusion of complex and diagnostically challenging cases but also justifying the need for biopsy in these patients. The considerable interobserver variability among dermatopathologists further prompted us to evaluate the added value of PsorX-LabDisk in particularly difficult cases. Therefore, we analyzed a subgroup of “ambiguous cases,” defined as those with <10 of 14 concordant diagnoses in the study cohort or disagreement among ≥1 of 3 dermatopathologists in the validation cohort. In total, 30 of 73 samples evaluated by dermatopathologists from the study cohort were classified as "ambiguous" (Fig. 3A), comprising 43.3% psoriasis and 56.7% eczema samples. Observer concordance compared to the reference diagnosis was markedly reduced, with roughly equal rates of correct and incorrect diagnoses for unclear samples (FPR = 37% ± 18.5%, FNR = 40% ± 22.3%, Fig. 3B). Expectedly, pairwise comparison confirmed higher interobserver variability than in the full dataset, with Cohen’s κ ranging from –0.28 to 0.37 in the study cohort (Fig. 3C) and –0.17 to 0.03 in the validation cohort (Fig. 3D). Accordingly, overall agreement for ambiguous samples was poor (Fleiss’ κ = 0.08 and –-0.11, respectively; Fig. 3E). We next aimed to assess diagnostic accuracy in dermatopathologically ambiguous cases by calculating sensitivity and specificity relative to the reference diagnosis. In the study cohort, sensitivity values varied between 0% to 83.3%, with an overall sensitivity of 61.4%. Specificity values were more consistent, ranging from 27.3% to 88.2% (mean specificity = 62%), corresponding to an overall diagnostic accuracy of 61.7% (Fig. 3F). In contrast, manual PCR and PsorX-LabDisk maintained high diagnostic performance even in these challenging cases, with identical parameters for sensitivity of 84.6%, specificity of 88.2 % and overall accuracy of 86.7% (Fig. 3G).

**Figure 3:**
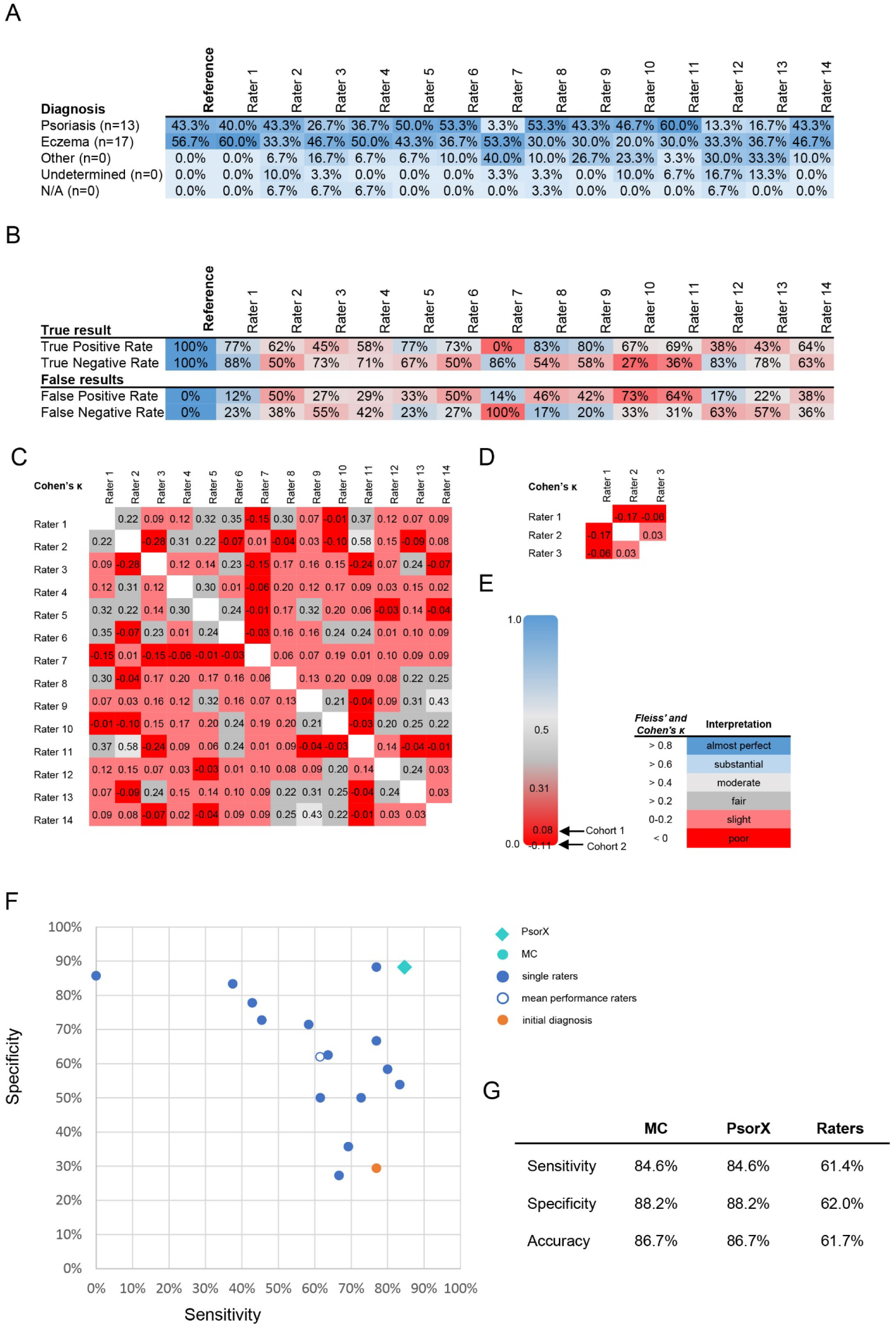
Interrater variability and diagnostic performance among dermatopathologists in ambiguous cases. **(A)** Distribution of diagnoses assigned by 14 dermatopathologists (Rater 1-14) compared to the reference diagnosis in the study cohort. **(B)** Diagnostic performance metrics for each rater. True positive rate (TPR) = proportion of psoriasis cases correctly identified as psoriasis; true negative rate (TNR) = proportion of eczema cases correctly identified as eczema; false positive rate (FPR) = proportion of eczema cases incorrectly diagnosed as psoriasis; false negative rate (FNR) = proportion of psoriasis cases incorrectly diagnosed as eczema. **(C–D)** Pairwise interrater agreement assessed using Cohen’s κ for the study cohort (C) and the validation cohort (D). **(E)** Agreement as measured by Fleiss’ κ and Cohen’s κ with interpretation according to Landis & Koch. **(F)** Sensitivity and specificity of individual dermatopathologists (blue dots), mean performance of all dermatopathologists (white dot outlined in blue), initial clinical diagnosis (orange), and molecular assays (MC, turquoise; PsorX, turquoise square) in the study cohort. **(G)** Summary of diagnostic performance metrics showing sensitivity, specificity, and overall accuracy for the manual molecular classifier (MC), fully automated PsorX-LabDisk system, and dermatopathology raters in the study cohort.

### Representative cases illustrate the diagnostic value of the molecular classifier in unclear cases

To further illustrate the potential of the molecular classifier to support diagnostic decision-making, three representative patient examples highlight the clinical and histological overlap underlying diagnostic ambiguity between psoriasis and eczema. Patient 1 was a male in his sixties who showed nummular psoriasiform plaques resembling both psoriasis vulgaris and nummular eczema. 6/14 dermatopathologists classified the lesion as eczema, while 8/14 voted for psoriasis. PsorX-LabDisk showed a clear probability for psoriasis (76.1%) and accordingly, the patient responded to TNF-inhibitor therapy (Fig. 4A-C). Another case with clinical suspicion for pustular palmoplantar psoriasis or superinfected dyshidrotic hand eczema yielded evenly split dermatopathological assessments between eczema and a completely other diagnosis. In line with the patient’s therapy response to TNF- and IL17 inhibition PsorX-LabDisk delivered a probability of 68.1% for the diagnosis of psoriasis (Fig. 4D-F). A third patient, showing psoriasiform skin lesions alongside psoriatic nail dystrophy, was predominantly interpreted as psoriasis (10/14 dermatopathologists). Interestingly, PsorX-LabDisk showed a probability of only 24.3% for psoriasis. In fact, the patient did not respond to TNF or IL17 inhibition and had in the meantime died of mycosis fungoides (Fig. 4G-I). Although PsorX-LabDisk was developed only to differentiate between eczema and psoriasis, a discrepancy between the PsorX-LabDisk result and the clinical and dermatopathological examination may indicate a need to re-examine the diagnosis.

**Figure 4:**
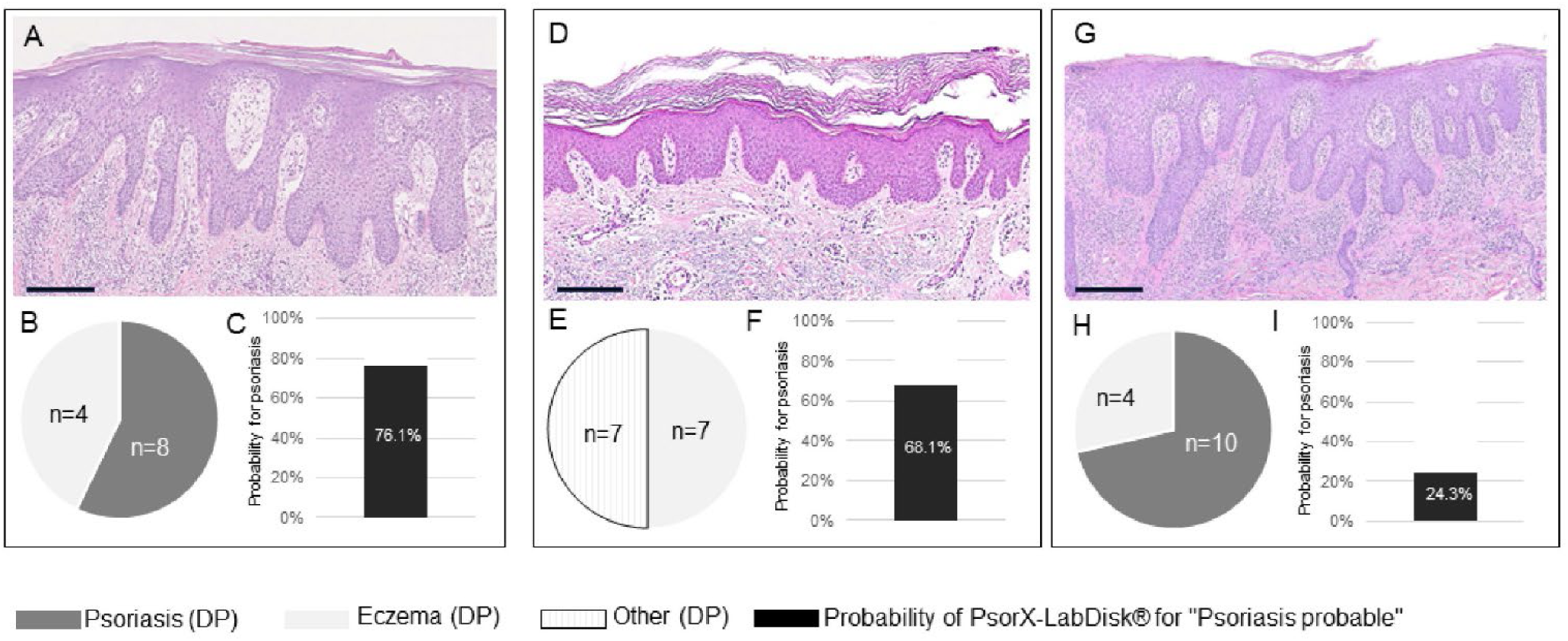
Representative cases illustrating diagnostic ambiguity between psoriasis and eczema and corresponding PsorX-LabDisk results. **(A–C)** Patient 1 presented with nummular psoriasiform plaques clinically suggestive of both psoriasis vulgaris and nummular eczema (Clinical images available upon request from the corresponding author) while histopathology showed psoriasiform hyperplasia (A). Eight of 14 dermatopathologists diagnosed psoriasis, while six favored eczema (B). PsorX-LabDisk indicated a 76.1% probability for psoriasis (C), consistent with therapeutic response to TNF inhibition. **(D-F)** Patient 2 presented with eczematous lesions at the elbow and palmar pustular lesions (Clinical images available upon request from the corresponding author) with rather eczematous features in histology (D). Diagnoses were evenly split between eczema and a diagnosis other than psoriasis or eczema (E). PsorX-LabDisk indicated a 68.1% probability for psoriasis (F), in line with clinical improvement under TNF- and IL-17-targeted therapy. **(G-I)** Patient 3 presented with psoriasiform plaques and nail dystrophy (Clinical images available upon request from the corresponding author) with dermatopathology showing psoriasiform changes (G). Ten of 14 dermatopathologists diagnosed psoriasis (H), whereas PsorX-LabDisk yielded a 24.3% probability for psoriasis (I). The patient did not respond to various psoriasis therapies and was later diagnosed with mycosis fungoides. Scale bars = 200 µm. DP = dermatopathologists.

## Discussion

The clinical and dermatopathological features of psoriasis and eczema frequently overlap, posing persistent challenges in differential diagnosis and leading to considerable rates of misclassification and diagnostic delay. Lauffer et al. reported that 15 % of psoriasis cases were initially diagnosed as eczematous dermatitis (*20*), and in palmoplantar disease, where phenotypic features often converge, misclassification may affect up to 50 % of cases (*21*). In primary care, psoriasis is also commonly mistaken for eczema or tinea, with diagnostic delays of up to five years (*22*). These challenges are further amplified in patients with skin of color: Brown et al. observed higher rates of misdiagnosis and delayed recognition of psoriasis among Hispanic and Black patients compared with White patients (*23*).

Consistent with these findings, our study showed that the initially suspected diagnosis differed from the final reference diagnosis in 34.3 % of cases in the study cohort. Given this clinical ambiguity, dermatopathological examination is routinely performed to support diagnostic decision-making. To evaluate the reproducibility of histopathological interpretation, lesions were independently reviewed by several dermatopathologists across two cohorts. Overall, interobserver agreement was only fair to moderate, reflecting the inherent subjectivity of dermatopathological assessment. In diagnostically ambiguous cases, concordance dropped markedly, emphasizing the limitations of morphology-based evaluation when inflammatory patterns overlap or remain subtle. This aligns with findings from a large multicenter study on melanocytic tumors, where full concordance among eight expert pathologists was achieved in only 53.5 % of cases, and for non-invasive melanomas in just 10 of 73 cases (*24*). These results highlight that dermatopathology which is often regarded as the diagnostic gold standard is itself subject to considerable interrater variability. Nevertheless, it needs to be emphasized that in our study only cases in which dermatologists considered a biopsy clinically necessary were included, resulting in a study population enriched for diagnostically ambiguous presentations. Consequently, interobserver agreement among dermatopathologists might be higher in more typical or unequivocal lesions. Moreover, inflammatory skin diseases generally require clinicopathological correlation, as no single histologic feature is sufficient for diagnosis. Moreover, the true diagnosis could only be assessed relative to the predefined reference (gold standard) diagnosis, which may itself be subject to inherent uncertainty.

Several molecular approaches have been proposed to complement traditional dermatopathological diagnosis and provide more objective discrimination between psoriasis and eczema. DaCosta et al. recently report IL-36γ immunohistochemical stainings as a reliable method to distinguish psoriasis from eczema with approximately 78.6% sensitivity in 25 samples (*25*), while Wittman et al. found that IL36γ combined with elafin serve as robust molecules distinguishing psoriasis and eczema-associated inflammation in epidermal tape strips (*26, 27*). The most established tool for discriminating psoriasis from eczema on molecular level has been introduced with a minimal gene-expression classifier based on *NOS2* and *CCL27*, first introduced in 2014, since then been validated across multiple cohorts (*17, 18, 28–30*) and recognized in reviews and guideline summaries as a leading classifier in this space (*31–33*). Surveys among members of the German Dermatologists’ Association (BVDD) (*34*) show growing interest in molecular diagnostics, which are currently used mainly for infectious diseases but increasingly seen as valuable for inflammatory skin disorders. In the 2024 BVDD yet unpublished survey with more than 200 participants, about 60 % of respondents viewed molecular testing as useful for unclear cases of psoriasis and eczema and over 80 % desired a test to differentiate psoriasis from eczema, while most emphasized that only fully automated solutions would be feasible in routine practice. This need has been addressed by PsorX-LabDisk, the first CE-IVD-certified test for differentiating psoriasis and eczema based on the proposed *NOS2/CCL27* classifier, enabling rapid and integrated PCR analysis from sample to answer in laboratory settings. In this study, we showed that not only the manual *NOS2/CCL27* classifier, but also PsorX-LabDisk, demonstrated superior diagnostic accuracy compared with the mean performance of dermatopathological evaluation, achieving a more favorable balance between sensitivity and specificity. Although certain individual observers reached similar values in selected parameters, the molecular assay yielded markedly higher reproducibility and consistent classification across both cohorts. Notably, in the subcohort of diagnostically ambiguous cases, both the manual *NOS2/CCL27*-based PCR classifier and the PsorX-LabDisk assay maintained high diagnostic accuracy, demonstrating robust performance even under challenging conditions.

Over the past two decades, highly specific biologics have revolutionized the treatment of psoriasis and eczema (*12*). However, their pathway-targeted mechanisms limit efficacy to defined patient subsets, emphasizing the need for precise diagnosis and therapy matching. In our study, the *NOS2/CCL27* classifier accurately reflected treatment response in patients with clinically and histologically ambiguous disease presentations. We also presented a patient case initially suspected to have psoriasis, in whom the *NOS2/CCL27* classifier indicated “psoriasis improbable.” This result was first interpreted as a false negative; however, several years later the patient was diagnosed with mycosis fungoides, suggesting that in some instances, the molecular classifier may reveal its diagnostic accuracy only in retrospect when the true disease becomes evident. As psoriasis and eczema represent only a fraction of the inflammatory disease spectrum, future classifiers must extend to broader differential diagnoses. Seremet et al. identified distinct immune gene expression modules across major inflammatory dermatoses, showing that such molecular signatures can refine diagnosis, enable patient stratification by immune pathway dominance, and support personalized, mechanism-based therapy selection. (*35*). These findings underscore the value of molecular diagnostics as an objective complement to dermatopathological evaluation, particularly in diagnostically challenging cases where expert interpretation may be uncertain. By reducing diagnostic ambiguity, such tools can enhance clinical confidence, enable more precise and cost-effective treatment decisions, and help avoid both delayed initiation of biologics and unnecessary exposure to systemic immunosuppressants.

## Data Availability

All data produced in the present study are available upon reasonable request to the authors.

## Abbreviations

AD: Atopic Dermatitis
EASI: Eczema Area and Severity Index
IL: Interleukin
PASI: Psoriasis Area and Severity Index
SCORAD: Scoring Atopic Dermatitis

## Data and materials availability

All data produced in the present study are available upon reasonable request to the authors.

## References

1. R. Parisi, D. P. Symmons, C. E. Griffiths, D. M. Ashcroft, Global epidemiology of psoriasis: a systematic review of incidence and prevalence. Journal of investigative dermatology 133, 377–385 (2013).

2. J. A. Odhiambo, H. C. Williams, T. O. Clayton, C. F. Robertson, M. I. Asher, I. P. T. S. Group, Global variations in prevalence of eczema symptoms in children from ISAAC Phase Three. Journal of Allergy and Clinical Immunology 124, 1251–1258.e1223 (2009).

3. J. I. Silverberg, Public health burden and epidemiology of atopic dermatitis. Dermatologic clinics 35, 283–289 (2017).

4. M. T. Fernandez-Figueras, L. Puig, Histopathological diagnosis of psoriasis and psoriasiform dermatitides. Diagnostic Histopathology 31, 87–97 (2025).

5. R. G. Phelps, M. K. Miller, F. Singh, The varieties of “eczema”: clinicopathologic correlation. Clinics in dermatology 21, 95–100 (2003).

6. M. Li, J. Wang, Q. Liu, Y. Liu, W. Mi, W. Li, J. Li, Beyond the dichotomy: understanding the overlap between atopic dermatitis and psoriasis. Frontiers in Immunology 16, 1541776 (2025).

7. J. M. Villa-Gonzalez, M. R. Gonzalez-Hermosa, J. Ugedo Alzaga, R. Pérez Blasco, P. Andrés Ibarrola, X. Atxutegi Ayesta, A. García García, I. Irizar Aguirre, B. Udondo González del Tánago, M. Pascual Ares, Interobserver Agreement of the Eczema Area and Severity Index for Atopic Dermatitis Severity Assessment: A Real-World Evidence Study. Dermatitis’, (2025).

8. C. Fink, C. Alt, L. Uhlmann, C. Klose, A. Enk, H. A. Haenssle, Intra-and interobserver variability of image-based PASI assessments in 120 patients suffering from plaque-type psoriasis. Journal of the European Academy of Dermatology and Venereology 32, 1314–1319 (2018).

9. A. Sprikkelman, R. Tupker, H. Burgerhof, J. Schouten, P. Brand, H. Heymans, W. Van Aalderen, A. Sprikkelman, Severity scoring of atopic dermatitis: a comparison of three scoring systems. Allergy 52, 944–949 (1997).

10. K. Gallagher, S. Halperin-Goldstein, A. S. Paller, New treatments in atopic dermatitis update. Ann Allergy Asthma Immunol, (2025).

11. A. W. Armstrong, A. Blauvelt, K. Callis Duffin, Y. H. Huang, L. J. Savage, L. Guo, J. F. Merola, Psoriasis. Nat Rev Dis Primers 11, 45 (2025).

12. A. Schabitz, K. Eyerich, N. Garzorz-Stark, So close, and yet so far away: The dichotomy of the specific immune response and inflammation in psoriasis and atopic dermatitis. J Intern Med, (2021).

13. P. Jonsson, A. C. Pilz, H. Maboudi, D. Ranzinger, P. Wagner, L.-N. Schaffert-Stone, C. Burg, M. S. Dadras, M. Bradley, F. Schauer, The Translational Dermatology Initiative– aiming at a new disease classification of inflammatory skin diseases. JID Innovations, 100381 (2025).

14. F. Fischer, A. Doll, D. Uereyener, S. Roenneberg, C. Hillig, L. Weber, V. Hackert, M. Meinel, A. Farnoud, P. Seiringer, J. Thomas, P. Anand, L. Graner, F. Schlenker, R. Zengerle, P. Jonsson, M. Jargosch, F. J. Theis, C. B. Schmidt-Weber, T. Biedermann, M. Howell, K. Reich, K. Eyerich, M. Menden, N. Garzorz-Stark, F. Lauffer, S. Eyerich, Gene expression based molecular test as diagnostic aid for the differential diagnosis of psoriasis and eczema in formalin fixed and paraffin embedded tissue, microbiopsies and tape strips. J Invest Dermatol, (2023).

15. A. M. D’Erme, D. Wilsmann-Theis, J. Wagenpfeil, M. Holzel, S. Ferring-Schmitt, S. Sternberg, M. Wittmann, B. Peters, A. Bosio, T. Bieber, J. Wenzel, IL-36gamma (IL-1F9) is a biomarker for psoriasis skin lesions. J Invest Dermatol 135, 1025–1032 (2015).

16. M. Della Mura, J. Sorino, A. Colagrande, M. Daruish, G. Ingravallo, A. Massaro, G. Cazzato, C. Lupo, N. Casatta, D. Ribatti, Artificial Intelligence in the Histopathological Assessment of Non-Neoplastic Skin Disorders: A Narrative Review with Future Perspectives. Medical Sciences 13, 70 (2025).

17. M. Quaranta, B. Knapp, N. Garzorz, M. Mattii, V. Pullabhatla, D. Pennino, C. Andres, C. Traidl-Hoffmann, A. Cavani, F. J. Theis, J. Ring, C. B. Schmidt-Weber, S. Eyerich, K. Eyerich, Intraindividual genome expression analysis reveals a specific molecular signature of psoriasis and eczema. Sci Transl Med 6, 244ra290 (2014).

18. N. Garzorz-Stark, L. Krause, F. Lauffer, A. Atenhan, J. Thomas, S. P. Stark, R. Franz, S. Weidinger, A. Balato, N. S. Mueller, F. J. Theis, J. Ring, C. B. Schmidt-Weber, T. Biedermann, S. Eyerich, K. Eyerich, A novel molecular disease classifier for psoriasis and eczema. Exp Dermatol 25, 767–774 (2016).

19. F. Fischer, A. Doll, D. Uereyener, S. Roenneberg, C. Hillig, L. Weber, V. Hackert, M. Meinel, A. Farnoud, P. Seiringer, J. Thomas, P. Anand, L. Graner, F. Schlenker, R. Zengerle, P. Jonsson, M. Jargosch, F. J. Theis, C. B. Schmidt-Weber, T. Biedermann, M. Howell, K. Reich, K. Eyerich, M. Menden, N. Garzorz-Stark, F. Lauffer, S. Eyerich, Gene Expression-Based Molecular Test as Diagnostic Aid for the Differential Diagnosis of Psoriasis and Eczema in Formalin-Fixed and Paraffin-Embedded Tissue, Microbiopsies, and Tape Strips. J Invest Dermatol, (2023).

20. F. Lauffer, K. Eyerich, Eczematized psoriasis - a frequent but often neglected variant of plaque psoriasis. J Dtsch Dermatol Ges 21, 445–453 (2023).

21. M. Kolesnik, I. Franke, A. Lux, S. R. Quist, H. P. Gollnick, Eczema in Psoriatico: An Important Differential Diagnosis Between Chronic Allergic Contact Dermatitis and Psoriasis in Palmoplantar Localization. Acta Derm Venereol 98, 50–58 (2018).

22. M. Abo-Tabik, R. Parisi, C. Morgan, S. Willis, C. E. Griffiths, D. M. Ashcroft, Mapping opportunities for the earlier diagnosis of psoriasis in primary care settings in the UK: results from two matched case–control studies. British Journal of General Practice, (2022).

23. S. G. Brown, C. B. Cobb, V. M. Harvey, Racial and ethnic health disparities in dermatology. Dermatologic Clinics 41, 325–333 (2023).

24. S. Haggenmüller, C. Wies, J. Abels, J. T. Winterstein, L. Heinlein, C. Nogueira Garcia, J. S. Utikal, S. A. Wohlfeil, F. Meier, S. Hobelsberger, Discordance, accuracy and reproducibility study of pathologists’ diagnosis of melanoma and melanocytic tumors. Nature Communications 16, 789 (2025).

25. A. DaCosta, G. H. Nguyen, T. S. Tran, T. M. Nguyen, M. T. McEvoy, D. A. Wetter, J. A. Vrana, A. Todd, A. C. Roden, J. S. Lehman, Differentiating psoriasis from eczema with interleukin-36 gamma immunohistochemistry on skin biopsy specimens: A retrospective, single-center validation and cohort study. Journal of the American Academy of Dermatology, (2025).

26. A. Berekmeri, A. Latzko, A. Alase, T. Macleod, J. S. Ainscough, P. Laws, M. Goodfield, A. Wright, P. Helliwell, S. Edward, G. D. Brown, D. M. Reid, J. Wenzel, M. Stacey, M. Wittmann, Detection of IL-36gamma through noninvasive tape stripping reliably discriminates psoriasis from atopic eczema. J Allergy Clin Immunol 142, 988–991 e984 (2018).

27. A. Berekmeri, T. Macleod, I. Hyde, G. J. Ojak, C. Mann, D. Kramer, M. Stacey, M. Wittmann, Epidermal proteomics demonstrates Elafin as a psoriasis-specific biomarker and highlights increased anti-inflammatory activity around psoriatic plaques. Journal of the European Academy of Dermatology and Venereology 39, 1324–1335 (2025).

28. P. M. Brunner, A. Israel, N. Zhang, A. Leonard, H. C. Wen, T. Huynh, G. Tran, S. Lyon, G. Rodriguez, S. Immaneni, A. Wagner, X. Zheng, Y. D. Estrada, H. Xu, J. G. Krueger, A. S. Paller, E. Guttman-Yassky, Early-onset pediatric atopic dermatitis is characterized by TH2/TH17/TH22-centered inflammation and lipid alterations. J Allergy Clin Immunol 141, 2094–2106 (2018).

29. P. Bentz, K. Eyerich, E. Weisshaar, Psoriasis or eczema? One-year results from the DGUV research project FB323 with application of the Molecular Classifier in occupational dermatoses. J Dtsch Dermatol Ges 20, 1233–1234 (2022).

30. F. Fischer, S. Roenneberg, L. Graner, F. Schlenker, R. Zengerle, F. J. Theis, C. B. Schmidt-Weber, T. Biedermann, F. Lauffer, N. Garzorz-Stark, S. Eyerich, Gene expression based molecular test proves clinical validity as diagnostic aid for the differential diagnosis of psoriasis and eczema in formalin fixed and paraffin embedded tissue. medRxiv, 2022.2005.2018.22275097 (2022).

31. Y. Renert-Yuval, J. P. Thyssen, R. Bissonnette, T. Bieber, K. Kabashima, D. Hijnen, E. Guttman-Yassky, Biomarkers in atopic dermatitis-a review on behalf of the International Eczema Council. J Allergy Clin Immunol 147, 1174–1190 e1171 (2021).

32. H. Dickel, A. Bauer, R. Brehler, V. Mahler, H. F. Merk, I. Neustädter, K. Strömer, T. Werfel, M. Worm, J. Geier, German S1 guideline: Contact dermatitis. JDDG: Journal der Deutschen Dermatologischen Gesellschaft 20, 712–734 (2022).

33. A. Bauer, R. Brans, R. Brehler, M. Büttner, H. Dickel, P. Elsner, M. Fartasch, C. Herzog, S. M. John, A. Köllner, S2k guideline diagnosis, prevention, and therapy of hand eczema. JDDG: Journal der Deutschen Dermatologischen Gesellschaft 21, 1054–1074 (2023).

34. L. Tizek, B. Schuster, C. Gebhardt, K. Reich, R. von Kiedrowski, T. Biedermann, K. Eyerich, A. Zink, N. Garzorz-Stark, Molecular diagnostics in dermatology: An online survey to study usage, obstacles and requirements in Germany. J Dtsch Dermatol Ges 20, 287–295 (2022).

35. T. Seremet, J. Di Domizio, A. Girardin, A. Yatim, R. Jenelten, F. Messina, F. Saidoune, C. Schlapbach, S. Bogiatzi, F. Minisini, Immune modules to guide diagnosis and personalized treatment of inflammatory skin diseases. Nature communications 15, 10688 (2024).

